# Individualised quantitative susceptibility mapping reveals abnormal hippocampal iron markers in acute mild traumatic brain injury

**DOI:** 10.1101/2025.04.28.25326522

**Authors:** Christi A. Essex, Mayan J. Bedggood, Jenna L. Merenstein, Catherine Morgan, Helen C. Murray, Samantha J. Holdsworth, Richard L. M. Faull, Patria Hume, Alice Theadom, Mangor Pedersen

**Affiliations:** Department of Psychology and Neuroscience, Auckland University of Technology, Auckland 0627, New Zealand; Brain Imaging and Analysis Center, Duke University Medical Center, Durham, NC 27710, United States; Center for Advanced MRI, The University of Auckland, Auckland 1023, New Zealand; School of Psychology and Centre for Brain Research, The University of Auckland, Auckland 1023, New Zealand; Center for Brain Research, The University of Auckland, Auckland 1023, New Zealand; Department of Anatomy and Medical Imaging, Faculty of Medical and Health Sciences, The University of Auckland, Auckland 1023, New Zealand; Mātai Medical Research Institute, Gisborne 4010, New Zealand; Sports Performance Research Institute New Zealand, Auckland University of Technology, Auckland 0627, New Zealand

**Keywords:** basal ganglia, brain iron, hippocampal subfields, individualised profiles, mild traumatic brain injury, quantitative susceptibility mapping

## Abstract

Quantitative susceptibility mapping (QSM) is an advanced post-processing technique of magnetic resonance imaging data that can be leveraged as a surrogate marker of iron accumulation in the brain following mild traumatic brain injury (mTBI). However, subtle tissue content changes characteristic of this complex injury may be lost to group-wise averaging when standard statistical models are employed. To provide more clinically- and individually-relevant information, z-tests can be used to build personalised profiles of positive susceptibility as a marker of abnormal iron homeostasis. Here, we mapped subject-specific deviations in iron-related positive susceptibility across 10 bilateral segmentations of the hippocampal subfields and 15 basal nuclei. The healthy normal susceptibility distribution for each region-of-interest (ROI) was derived from the aggregate data of 25 age-matched male controls (*M* = 21.10 years [range: 16-32], *SD* = 4.35) using z-tests. Region-wise z-scores for each of the 35 males aged between 16 and 33 years (*M* = 21.60, *SD* = 4.98) with acute (< 14 days) sports-related mTBI (sr-mTBI) were compared against the healthy reference range. Of the sr-mTBI participants, 43% exhibited abnormal iron markers in at least one ROI, which involved the hippocampal subfields in a majority (87%) of cases. Across all ROIs, particularly dense concentrations were observed in the parasubiculum and mammillary nucleus. Injury severity scores were not significantly different between sr-mTBI participants with abnormal iron markers (*M* = 41.7, *SD* = 34.5) and those without (*M* = 35.6, *SD* = 30.8), *p* = 0.5, however, abnormal iron markers in certain hippocampal subfields and the mammillary nucleus were observationally linked to clinical symptom phenotype. Taken together, these data allude to a region-of-risk model in which areas of the anteromedial hippocampal head, which is proximal to the sphenoid ridge, and midline structures are vulnerable to iron-mediated pathology. These findings underscore the importance of subject-specific analyses and how these sensitive methods can be used to map regional iron dyshomeostasis against cranial-dural morphology and established injury biomechanics.

## Introduction

As the most prevalent neurological condition, traumatic brain injury (TBI) represents a significant global health concern.^1^ Projections estimate that TBI will remain one of the three foremost causes of injury-related death and disability into 2030.^2^ However, far from a binary condition, TBI exists along a continuum from mild to severe, with mild TBI (mTBI) accounting for the vast majority (∼90%) of all cases.^1^ Unlike severe TBI where lesions are readily identifiable on conventional clinical imaging, such as computed tomography (CT) and magnetic resonance imaging (MRI), the degree of tissue damage is comparatively less extreme for mTBI and standard radiographic methods fail to detect mTBI-related brain alterations in 90-95% of cases.^1^ As a result, mTBI diagnosis and prognosis relies almost exclusively on clinical assessment by subjective self-report,^3^ introducing a level of ambiguity that has led not only to definitional and diagnostic inconsistencies, but drastic inequities in access to care.^4, 5^ Here, the identification of objective markers sensitive to the inter-individual heterogeneity of micropathology is critical to advancing the current understanding of mTBI and facilitating targeted, equitable, interventions.

Iron, the most abundant trace metal in the brain, has been implicated in the secondary neurometabolic cascade^6–8^ and is associated with cognitive impairments following mTBI,^7, 9, 10^ highlighting its promise as a potential mTBI biomarker. Whilst the precise mechanisms by which excess iron accumulates in the brain is an area of ongoing research, injury-induced microvascular dysfunction, microbleeds, and increased blood-brain barrier (BBB) permeability likely increase iron transport into neural tissue.^6, 7, 11, 12^ Normally sequestered in ferritin to mitigate oxidative stress, supra-physiological levels of iron can overwhelm the capacity of storage proteins, resulting in a labile iron pool which catalyses highly cytotoxic redox reactions.^13, 14^ The reactive species generated by redox cycling precipitates oxidative stress, lipid peroxidation, increased cell membrane permeability, increased expression of pro-inflammatory cytokines, demyelination, damage to DNA, RNA, carbohydrates, lipids, and proteins, mitochondrial dysfunction, glutamate excitotoxicity, proteinopathy, and iron-regulated cell death.^6, 8, 11, 13–19^

Quantitative susceptibility mapping (QSM), an advanced MRI post-processing technique, leverages the magnetic field perturbations induced by biomagnetic substrates, including non-heme iron, to generate scalar maps for an indirect marker of tissue dyshomeostasis.^20–22^ While QSM has been extensively used to characterise iron dyshomeostasis in neurodegenerative disorders,^23–28^ this technique remains under-utilised in the context of mTBI.^29^ In the relatively small number of QSM investigations of mTBI, the total deep grey matter or major basal ganglia substructures including the thalamus, caudate, putamen, and globus pallidus remain the most commonly targeted subcortical regions of interest (ROIs) due to their high iron load in the brain.^30–36^ This high regional iron content also render the basal ganglia particularly amenable to imaging methods sensitive to tissue magnetic susceptibility, such as QSM. While this highlights the prevailing rationale for selecting the basal ganglia as logical targets in QSM-based mTBI research, the limited anatomical specificity may account for the lack of significant results indicative of iron accumulation in the subcortical grey matter.^30–36^ Even the two more extensive investigations inclusive of nucleus accumbens and/or amygdala segmentations^34, 36^ have not differentiated the subdivisions of basal ganglia, for example the internal and external segments of the globus pallidus, nor explored mTBI effects in other associated nuclei. Evidence from susceptibility-weighted imaging (SWI) identifying the substantia nigra and red nucleus as loci of iron dyshomeostasis post-mTBI^9^ highlights why exploration of additional structures may enable better characterisation of altered iron signalling in the brain after injury.

Additionally, despite the well-documented vulnerability of the hippocampus to injury,^37, 38^ inflammatory processes, impaired long-term potentiation,^39, 40^ increased expression of amyloid precursor protein,^41^ and a high expression of transferrin receptor protein 1 (TfR1) on the apical BBB endothelium^42^ which may exacerbate abnormal iron transport across this barrier following injury, investigations of this a crucial memory hub^43^ remain limited. Of the three extant studies to include the hippocampus as a region-of-interest (ROI),^34, 36, 44^ only our previous work^44^ has differentiated the hippocampal subfields. Here, we posited that limited anatomical specificity and treatment of the hippocampus as a homogeneous ROI may mask the iron-related pathology that has been detected by SWI^9^ but is absent from the findings of prior QSM-based research. Using an extensive 16-part segmentation of the basal nuclei and 10 hippocampal subfields, we identified decreased negative susceptibility in the cornu ammonis (CA) 4 region consistent demyelination and the selective vulnerability of hilar cell populations to injury.^45–49^ However, like prior investigations, we observed no significant increase in positive susceptibility as a marker of iron dyshomeostasis in deep grey matter.

Here, recent research highlighting the utility of subject-specific modelling to capture data that are otherwise lost to group-wise averaging^50–56^ is particularly relevant. The vast majority of clinical research employ analytic techniques that assess group averages and treat individual variability primarily as noise.^57^ These methods are used in an attempt to identify signatures representative of the “average patient”, and while more standard group-wise statistical approaches have been fundamental identifying disorder-specific neural signatures, such as structural alterations in the amygdala and hippocampus in anxiety^58^ and altered rostral anterior cingulate activity in depression,^59^ the generalisability at the individual level is limited.^57^ These macroscopic approaches likely obscure the individual-level heterogeneity that characterises clinical cohorts; variation that is central to identifying appropriate and effective intervention.^57^ Group-level analytic strategies remain the most common method used in QSM investigations of mTBI. The potential loss of subtle tissue responses to averaging effects could plausibly explain the general lack of significant effects across the extant literature. Indeed, we have previously demonstrated the capacity of individualised investigations to identify individual-level cortical iron markers ^60^ that are not apparent at the group level.^61^ In this individualised study, identification of a significantly higher symptom burden for mTBI participants presenting with abnormal iron markers relative to those exhibiting no evidence of iron dyshomeostasis further underscores the clinical relevance of more sensitive modelling. In the present investigation, we extend this individualised technique to deep grey matter, using the results to facilitate targeted analyses of differences in injury severity scores between mTBI participants with normal and abnormal subcortical iron markers as a secondary measure. Given the novel and exploratory nature of this study, no specific *a priori* hypotheses were made regarding the direction of effects across all basal nuclei and hippocampal subfields.

## Materials and Methods

Ethical approval for this research was obtained from the Health and Disabilities Ethics Committee (HDEC) (Date: 18/02/2022, Reference: 2022 EXP 11078) and institutional approval was also obtained from the Auckland University of Technology Ethics Committee (AUTEC) (Date: 18/02/2022, Reference: 22/12). In accordance with the Declaration of Helsinki, all participants provided written informed consent prior to data collection.

### Participants

Thirty-five male contact sports players (*M* = 21.60 years [range: 16-33], *SD* = 4.98) with acute sports-related mTBI (sr-mTBI) sustained within 14 days of MRI scanning (*M* = 10.40 days, *SD* = 3.01) and 25 age-matched male controls (*M* = 21.10 years [range: 16-32], *SD* = 4.35) were recruited for this observational study (see Table 1). To minimise confounds related to iron accumulation across the lifespan,^62–68^ we ensured ages were not significantly different between groups (*t*(58) = -0.44, *p* = 0.66). Clinical (sr-mTBI) participants were recruited through three Axis Sports Medicine clinics (Auckland, New Zealand), via print and social media advertisements, word-of-mouth, and through community-based pathways including referrals from healthcare professionals and sports team management. Each clinical participant was required to have a confirmed sr-mTBI diagnosis by a licensed physician as a prerequisite for study inclusion, and symptom severity was assessed using the Brain Injury Screening Tool (BIST)^69^ either upon presentation to Axis clinics or electronically following recruitment. Healthy controls (HC) were recruited through print and social media advertisements, and word-of-mouth. Exclusion criteria for all participants included a history of significant medical or neurological conditions unrelated to the study’s objectives and contraindications for MRI. Additionally, controls were excluded if they had any recent history of mTBI events (< 12 months) or were living with any long-term effects of previous mTBI. All participants completed a brief demographic questionnaire and attended a 1-hour MRI scan at The Centre for Advanced MRI (CAMRI), Auckland, New Zealand. All scans were reviewed by a certified neuroradiologist consultant for clinically significant findings. No clinically significant diagnoses were identified from MRI in either group, and no incidental findings required follow-up (see Table 1).

**Table 1:** Clinical characteristics of participants with mTBI.

| ID | Age | DSI | BIST Score | MOI | MRI findings |
| --- | --- | --- | --- | --- | --- |
| mTBI-01 | < 20 | 5 days | 140 | Rugby | None |
| mTBI-02 | < 20 | 5 days | 12 | Rugby | None |
| mTBI-03 | 20s | 6 days | 78 | Rugby | None |
| mTBI-04 | < 20 | 13 days | 18 | Rugby | Small fluid signal spaces in R peritrigonal WM - normal. R caudate cleft along ventricular surface - possibly developmental or from old ischaemic insult |
| mTBI-05 | < 20 | 12 days | 12 | Rugby | None |
| mTBI-06 | 20s | 13 days | 42 | Rugby | None |
| mTBI-07 | 20s | 13 days | 13 | Football | Minor artefactual T1 signal in pons |
| mTBI-08 | 20s | 12 days | 6 | Hockey | None |
| mTBI-09 | 20s | 6 days | 56 | Rugby | Minor R orbital fracture (old) |
| mTBI-10 | < 20 | 12 days | 54 | Rugby | None |
| mTBI-11 | 20s | 10 days | 52 | Rugby | None |
| mTBI-12 | 30s | 13 days | 13 | Football | None |
| mTBI-13 | < 20 | 5 days | 79 | Rugby | None |
| mTBI-14 | 20s | 13 days | 2 | Rugby | Small focus of susceptibility in L superior frontal gyrus possibly vascular or nonspecific haemosiderin |
| mTBI-15 | < 20 | 13 days | 22 | Rugby | None |
| mTBI-16 | < 20 | 8 days | 117 | Futsal | Tiny cleft of fluid signal in R cingulate gyrus - minor developmental anomaly or mature gliosis |
| mTBI-17 | 20s | 13 days | * | Rugby | None |
| mTBI-18 | 20s | 10 days | 34 | Gymnastics | None |
| mTBI-19 | 20s | 13 days | 28 | Jiu-jitsu | Some artefactual DWI signal in pons |
| mTBI-20 | 20s | 11 days | 69 | Surfing | Tiny susceptibility site in R temporal lobe - may be vascular |
| mTBI-21 | < 20 | 7 days | 14 | Rugby | Minor susceptibility in transverse sulcus in R mid temporal lobe - nonspecific, may be vascular or reflect haemosiderin deposition from prior small volume haemorrhage |
| mTBI-22 | < 20 | 13 days | 39 | Judo | None |
| mTBI-23 | < 20 | 9 days | 34 | Rugby | None |
| mTBI-24 | < 20 | 12 days | 68 | Rugby | None |
| mTBI-25 | 20s | 12 days | 17 | Rugby | 7mm pineal cyst - normal limits. Some T1 hyperintensity in R cerebellum - artefact compatible |
| mTBI-26 | 20s | 12 days | 25 | Rugby | Mildly prominent cisterna magna |
| mTBI-27 | 30s | 7 days | 30 | Football | A few mildly prominent biparietal and L cerebral peduncle perivascular spaces - normal variant |
| mTBI-28 | 30s | 12 days | 51 | Swimming | None |
| mTBI-29 | 20s | 5 days | 6 | Rugby | None |
| mTBI-30 | < 20 | 12 days | 2 | Rugby | Some DWI signal disturbance anterior to pons - likely artefactual |
| mTBI-31 | < 20 | 14 days | 22 | Rugby | 2-3 tiny foci of susceptibility in R frontal lobe - nonspecific, possible site of prior microhemorrhage. A punctate focus of T1 hypointensity/T2 hyperintensity superolateral to the frontal horn of R lateral ventricle |
| mTBI-32 | 20s | 8 days | 58 | Football | Bifrontal developmental venous anomaly noted - normal variants |
| mTBI-33 | < 20 | 8 days | 8 | Rugby | Minuscule foci of susceptibility in R cerebellar hemisphere /posterior to R aspect of the splenium of CC - nonspecific. Minor susceptibility in R sylvian fissure - vascular |
| mTBI-34 | 20s | 14 days | 47 | Rugby | None |
| mTBI-35 | < 20 | 12 days | 28 | Football | None |
| **Mean mTBI** | 21.60 (4.98) years | 10.4 (3.01) days | 38.1 (32.0) /160 |  | No findings considered clinically relevant |
| **Mean HC** | 21.10 (4.35) years |  |  |  | No findings considered clinically relevant |

*Note.* Diagnostic assessment is limited to the volume T1, SWI and DWI sequences with only limited interpretation of the multi-echo T2 stack. Clinical assessments are relevant to the identification of micro-haemorrhages, areas of siderosis, T1 appearance, gliosis, volume, ventricular volumes and non-neurological findings. Age is given in a range to prevent re-identification of participants. The possible range of BIST scores is 0 (min) to 160 (max). Clinical group data correspondent to date at MRI only with the exception of the BIST<sup>69</sup> acquired >24 hours post-injury prior to MRI scanning (<14 days post). Abbreviations are as follows: mTBI = mild traumatic brain injury; HC = healthy control; ID = unique identifier; DSI = days since injury; BIST = Brain Injury Screening Tool; MOI = mechanism of injury; MRI = magnetic resonance imaging; L = Left; R = right; \* = missing data (BIST incomplete on the Axis Sport Medicine Clinic patient portal, reason unknown).

### Neuroimaging

Details on image acquisition and processing have been previously reported,^44, 61^ and are summarised here for brevity.

### Acquisition

MRI data were acquired on a 3T Siemens MAGNETOM Vida Fit scanner (Siemens Healthcare, Erlangen, Germany) equipped with a 20-channel head coil. A 3D flow-compensated single-echo gradient-recalled echo (GRE) sequence was used to obtain magnitude and unfiltered phase images for QSM reconstruction. Data were collected at 1 mm isotropic voxel size with matrix size = 180 x 224 x 160 mm, repetition time (TR) = 30 ms; echo time (TE) = 20 ms; flip angle (FA) = 15*^◦^*; field of view (FoV) = 180 mm (LR) × 224 mm (AP) in a total acquisition time of ∼3.43 minutes. For each participant, a high-resolution 3D *T*_1_-weighted (*T*_1_w) anatomical image volume was acquired for coregistration and segmentation using a Magnetisation-Prepared Rapid Acquisition Gradient Echo (MPRAGE) sequence (TR = 1940.0 ms; TE = 2.49 ms, FA = 9*^◦^*; slice thickness = 0.9 mm; FoV = 230 mm; matrix size = 192 x 512 x 512 mm; GRAPPA = 2; voxel size 0.45 x 0.45 x 0.90 mm) for a total acquisition time of ∼4.31 minutes.

### QSM processing

QSM images were reconstructed using the QSMxT^70^ v6.4.2 pipeline with two-pass artefact reduction.^71^ The rapid open-source minimum spanning tree algorithm algorithm was used for phase unwrapping,^72^ background field removal was performed using projection onto dipole fields,^73^ and rapid two-step dipole inversion was used for susceptibility estimation.^74^ Susceptibility was referenced to the whole-brain in accordance with recent consensus statement recommendations.^75^

Magnitude images were skull-stripped with FSL’s *BET*^76^ and used to generate binary masks for signal erosion at the brain perimeter on QSM using *fslmaths*. Skull-stripped magnitude images were linearly registered to the CIT168 *T*_1_w template^77^ with 12 degrees of freedom (DoF) suitable for atlas-based registration using FMRIB’s Linear Transformation Tool.^78–80^ The corresponding transformation matrix was used for spatial normalisation of the QSM images using FLIRT.^78–80^ QSM images were then thresholded above and below zero to generate separate maps of net positive (iron-related) and net negative (myelin-, calcium-, and protein-related) voxel-wise susceptibility.^81, 82^ Only net positive susceptibility maps (*QSM* ^+^) were retained for statistical analysis of iron markers.

### Basal ganglia segmentation

The 16-part CIT168 bilateral basal ganglia mask^77^ in MNI152 space was used for segmentation of the putamen (Pu), caudate (Ca), nucleus accumbens (NAC), globus pallidus externus (GPe), globus pallidus internus (GPi), ventral pallidum (VeP), substantia nigra pars compacta (SNc), substantia nigra pars reticulata (SNr), extended amygdala (EXA), hypothalamus (HTH), mammillary nucleus (MN), ventral tegmental area (VTA), parabrachial pigmented nucleus (PBP), habenular nuclei (HN), subthalamic nucleus (STH), and the red nucleus (RN).

### Hippocampal segmentation

Each participants’ *T*_1_w structural image was processed using FreeSurfer’s *recon-all* pipeline^83^ to generate a 10-part unilateral segmentation of the hippocampal subfields.^84^ The hippocampal subregion masks include the parasubiculum, presubiculum, subiculum, cornu ammonis (CA) regions CA1, CA3 (which includes CA2), and CA4, the hippocampal-amygdala transition area (HATA), fimbria, hippocampal tail, and hippocampal fissure. To standardise each hippocampal subfield mask for data analysis, each participants’ *T*_1_w brain image was registered to the CIT168 *T*_1_w template in MNI152 space^77^ with 12 DoF using FLIRT.^78–80^ The resulting transformation matrix was then applied to both the left and right hemisphere hippocampal masks using nearest-neighbour interpolation.

### Personalised QSM profiles

Individual profiles of positive susceptibility were generated at the bilateral level using MATLAB (2024a). Mean positive susceptibility values were extracted from each of the 16 bilateral basal ROIs and 10 unilateral hippocampal segmentations. To yield a bilateral measure, left and right hemisphere susceptibility values were averaged for each hippocampal subfield. The HN was omitted due to zero values in this ROI for a subset of participants. No zero values were present for any participant within any hippocampal ROI. Z-scores were calculated for all participants (HC and mTBI), by subtracting the HC group mean from each individual’s susceptibility value and dividing by the HC group standard deviation in accordance with methods used across different imaging modalities to investigate the effects of TBI and mTBI.^50–55, 60^ To bring the HC data closer to a normal distribution, outlier scores for the HC group were filtered if they fell outside 2 times the interquartile range (IQR), a more stringent criterion than the methods used to identify mild outliers 1.5 times the IQR, but less extreme than the more conservative filter of 3 times the IQR.^85^ HC outliers were filtered in the hippocampal parasubiculum (*n* = 2), presubiculum (*n* = 2), and subiculum (*n* = 1). For basal ROIs, *n = 1* HC participant was excluded in the EXA and VeP, *n = 2* participants were excluded in the STH, and *n = 6* participants were excluded in the MN. Despite the more extensive filtering applied to the MN region, the reduced HC sample size for this region (*n* = 19) is larger than comparable individualised QSM studies investigating moderate-to-severe TBI that have used a sample size of 12.^53^ After filtering, the Shapiro-Wilk normality test yielded an average *W* value of *M* = 0.95 (*SD* = 0.02) across hippocampal ROIs and an average *W* value of *M* = 0.94 (*SD* = 0.03) across basal nuclei, indicating that the data distribution within each ROI was close to normal. The pipeline is summarised in Fig. 1 below.

**Fig 1:**
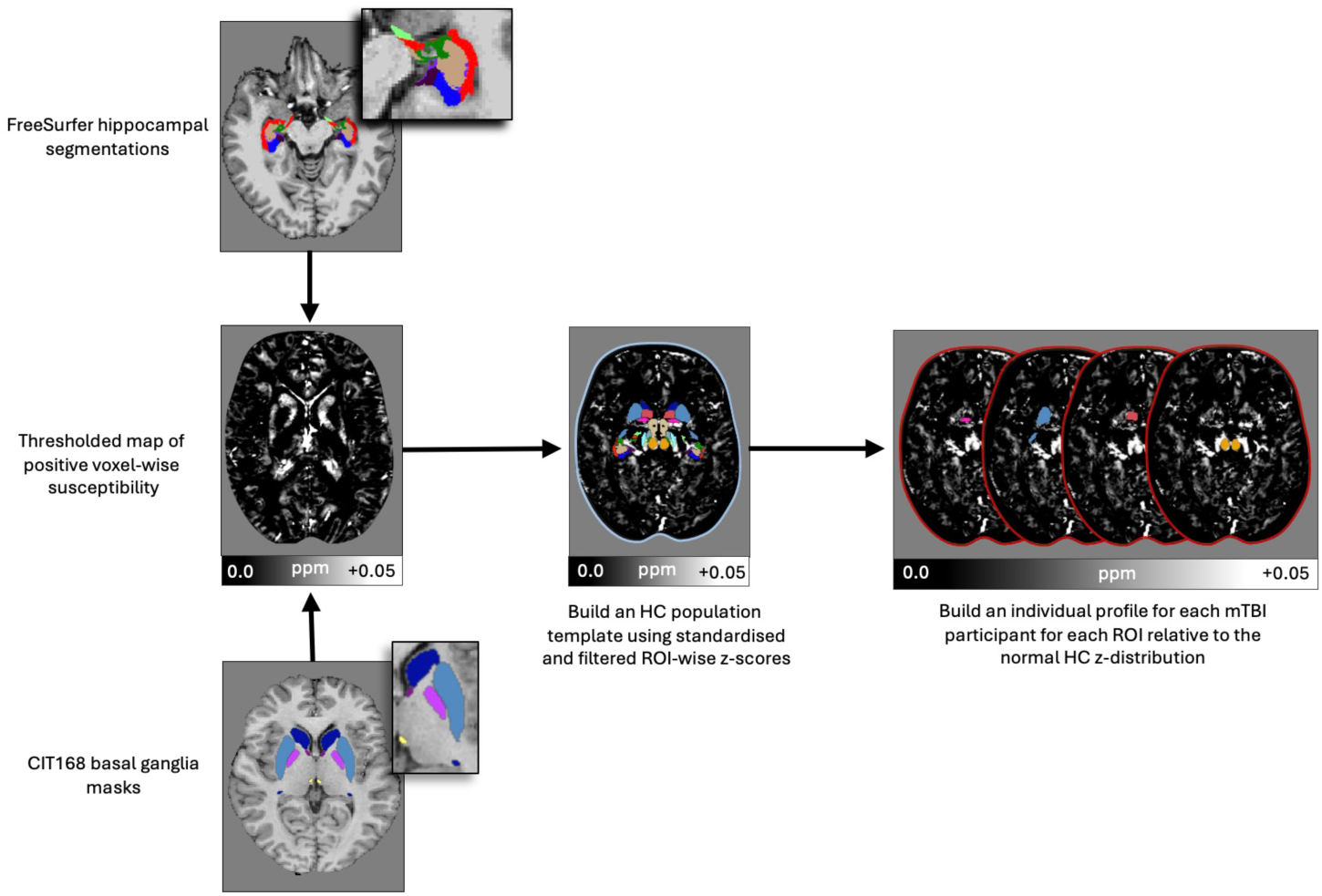
Pipeline for individual subcortical iron profile generation. FreeSurfer was used to generate segmentations of the hippocampal subfields from each participants’ *T*_1_w image volume, and the CIT168-MNI152 basal ganglia mask was used to delineate subcortical nuclei. These segmentations are overlaid on a structural *T*_1_w image volume for enhanced visualisation (top left = hippocampal segmentations; bottom left = basal ganglia masks). QSM images were thresholded for net positive susceptibility (*QSM* ^+^ = centre left image). After voxel-wise thresholding, mean positive susceptibility values were extracted for each hippocampal and basal ROI. Z-scores were calculated using the mean and standard deviation of the healthy control group, and standardised around a mean of zero. The HC distribution was filtered to remove outliers exceeding two times IQR for a normal distribution. Each mTBI participants’ z-score were compared to the healthy ROI-wise normal distribution, while controlling for multiple comparisons. QSM = quantitative susceptibility mapping; ROI = region of interest; mTBI = mild traumatic brain injury; HC = healthy control; IQR = interquartile range.

### Statistical analyses

To assess statistical significance for mTBI z-scores, two-tailed p-values were calculated from the z-scores using the cumulative distribution function of the standard normal distribution. A false discovery rate (FDR) correction^86^ was separately applied to the p-values for the 10 hippocampal or 15 basal nuclei ROI-wise comparisons.

To conduct secondary exploratory statistical tests, the mTBI group was divided into two subgroups: those whose z-scores significantly deviated from the HC normal distribution for any hippocampal or basal ROI (i.e., iron-abnormal) and those whose scores did not (i.e., iron-normal). Although preliminary analyses indicated no statistically significant difference in age between mTBI and HC participant groups, an ANOVA was performed between these three new subgroups (iron-normal, iron-abnormal, and controls) to confirm that age was not driving the identification of abnormal z-scores. Iron deposition in subcortical nuclei is a recognised feature of healthy ageing, particularly in individuals of a similar age range to this cohort,^62, 65, 66^ thereby necessitating this further analysis to ensure that observed results were not confounded by normative, age-related tissue changes. Additionally, a nonparametric Mann-Whitney U test was used to assess whether injury severity (BIST scores) differed significantly between iron-abnormal and iron-normal mTBI participants, excluding mTBI-17 for this analysis only due to missing BIST data (see Table 1).

## Results

### Regional individualised subcortical iron profiles

Of the 35 total mTBI participants, 15 (43%) exhibited significantly higher z-scores than the healthy population template in at least one ROI (see Table 2, Fig. 2.2A and Fig. 2.2B). Among these “iron-abnormal” participants, 87% expressed abnormal iron markers inclusive of hippocampal ROIs, which frequently involved the parasubiculum. In contrast, only 60% of participants had abnormal iron markers inclusive of basal ganglia subregions. These abnormal markers were characterised by a relatively narrow distribution of z-scores across the available basal ganglia segmentations, with abnormal z-scores most commonly observed in the mammillary nucleus.

**Fig 2:**
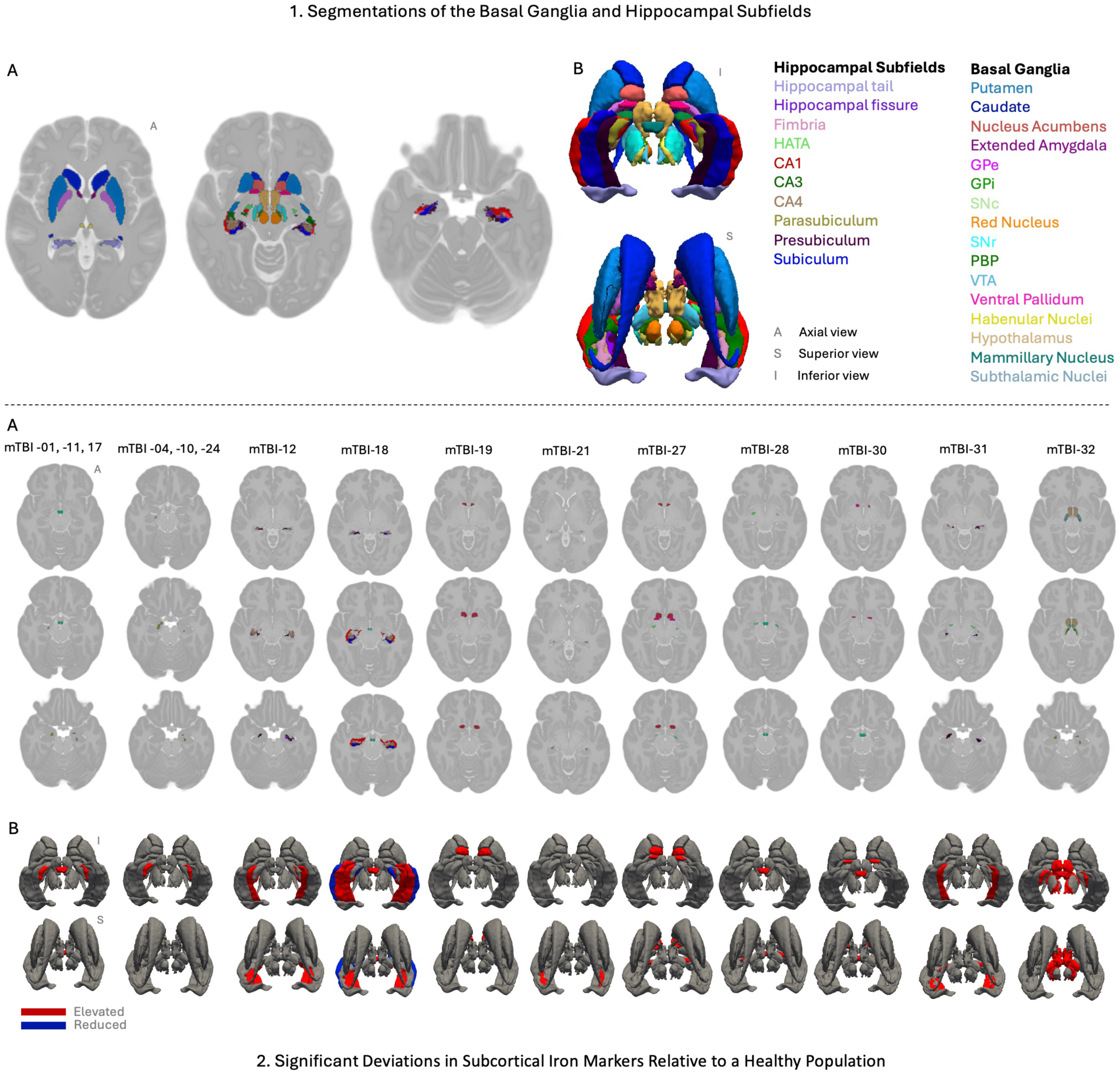
Segmentations and individualised profiles for abnormal subcortical iron markers. 2. Visualisation of basal ganglia and hippocampal subfield segmentations. A. Axial (A) slices of the CIT168-MNI152 *T*_1_w brain template depicting segmentations of the basal ganglia and hippocampal subfields, selected to optimise structural visualisation. B. 3D renderings show superior (S) and inferior (I) views of the segmented regions, with key subcortical structures labelled in their respective colours. 2. Significant deviations in subcortical iron markers for each sr-mTBI participant relative to the healthy control population. A. Axial views of the CIT168-MNI152 *T*_1_w template highlight significant subcortical ROIs for each participant, shown in the segmentation colours. B. 3D renderings highlight regions with significantly elevated (red) or reduced (blue) z-scores to show regions with abnormal iron profiles; all other regions are greyed out. CA = cornu ammonis; GP = globus pallidus (GPi = internus; GPe = externus); SN = substantia nigra (SNc = pars compacta; SNr = pars reticulata); PBP = parabrachial pigmented nucleus; VTA = ventral tegmental area.

**Table 2:**
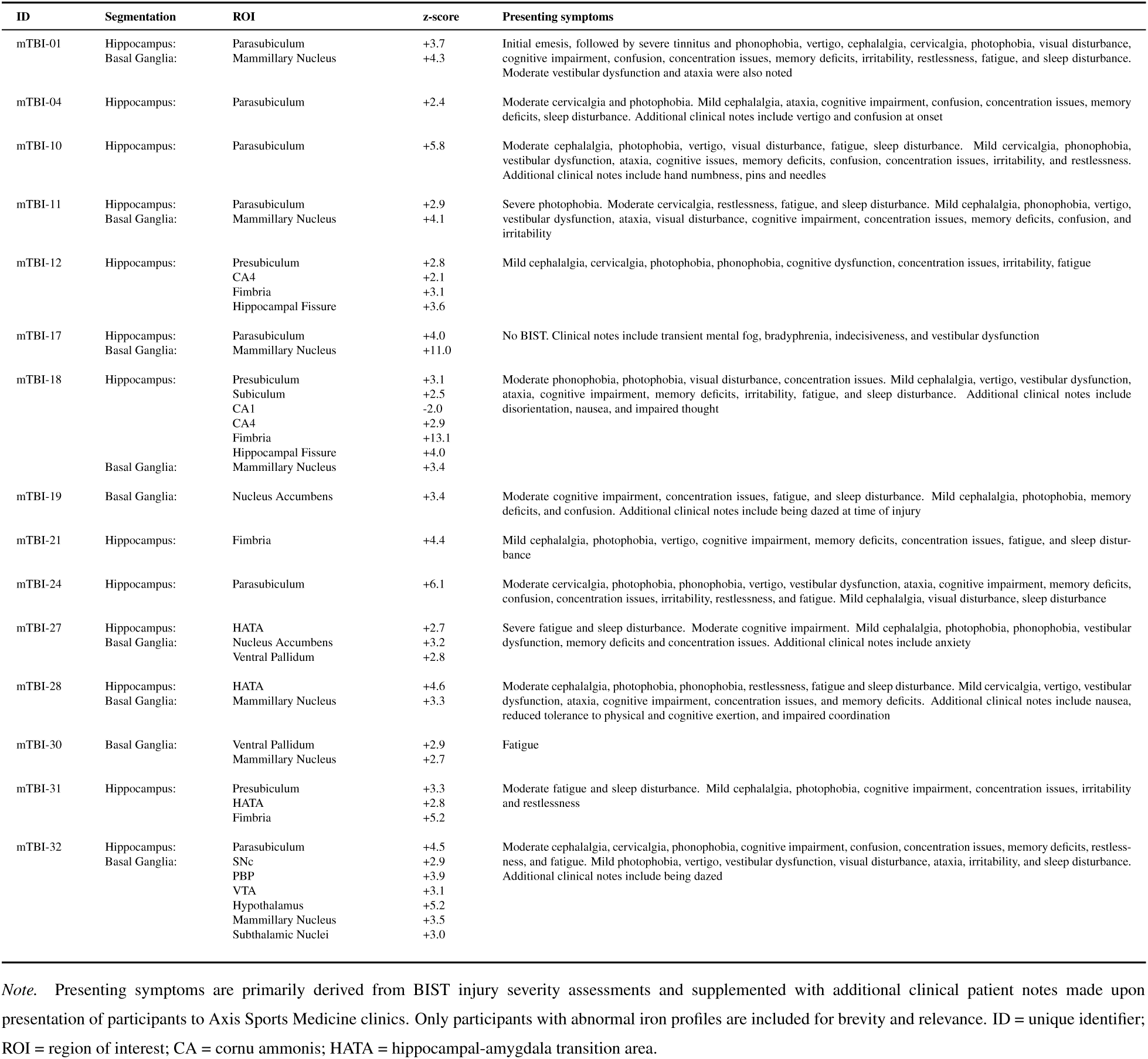
Summary of significant ROI-wise z-scores and symptomatology for iron-abnormal sr-mTBI participants in the basal ganglia and hippocampal subfields.

In addition to the high incidence of elevated z-scores in the hippocampal parasubiculum and basal mammillary nucleus, both of which were affected in approximately half of the 15 participants with abnormal iron markers (47%), relatively lower incidences of iron-related abnormalities were apparent in the hippocampal fimbria (27%), presubiculum and HATA (20% each) (see Fig. 2.2A and Fig. 2.2B). Less frequently affected ROIs included the hippocampal CA4 region and hippocampal fissure, as well as the basal nucleus accumbens and ventral pallidum, each with a 13% incidence rate. The basal SNc, PBP, hypothalamus, VTA, subthalamic nucleus, and hippocampal subiculum exhibited only isolated instances of elevated susceptibility across the whole iron-abnormal cohort. Only two hippocampal ROIs exhibited no significant deviations from the healthy normal distribution for any mTBI participant; the hippocampal tail and CA3. In contrast, no abnormal iron profiles were identified for several major basal subregions, including the putamen, caudate, globus pallidus (both internus and externus), red nucleus, SNr, and extended amygdala.

Only one participant (mTBI-18) had a z-score below the healthy control range, showing a *z* of -2.0 in the CA1 region. This case also represents the sole instance of CA1 involvement (see Table 2, Fig. 2.2A and Fig. 2.2B).

On aggregate, incidence rates of abnormal iron markers tended to cluster around the midline, in commissural nuclei, and inferior regions (see Fig. 3). A particularly high concentration was observed in the hippocampus, which is located inferior to the basal ganglia. Within the hippocampus, prevalent abnormal iron markers were observed near the anteromedial aspect of the hippocampus (the hippocampal head), decreasing in frequency laterally and toward the distal tail. In contrast, more superior regions, including the dorsal aspect of the hippocampus and the basal ganglia (excluding the mammillary nucleus) appeared to be less affected.

**Fig 3:**
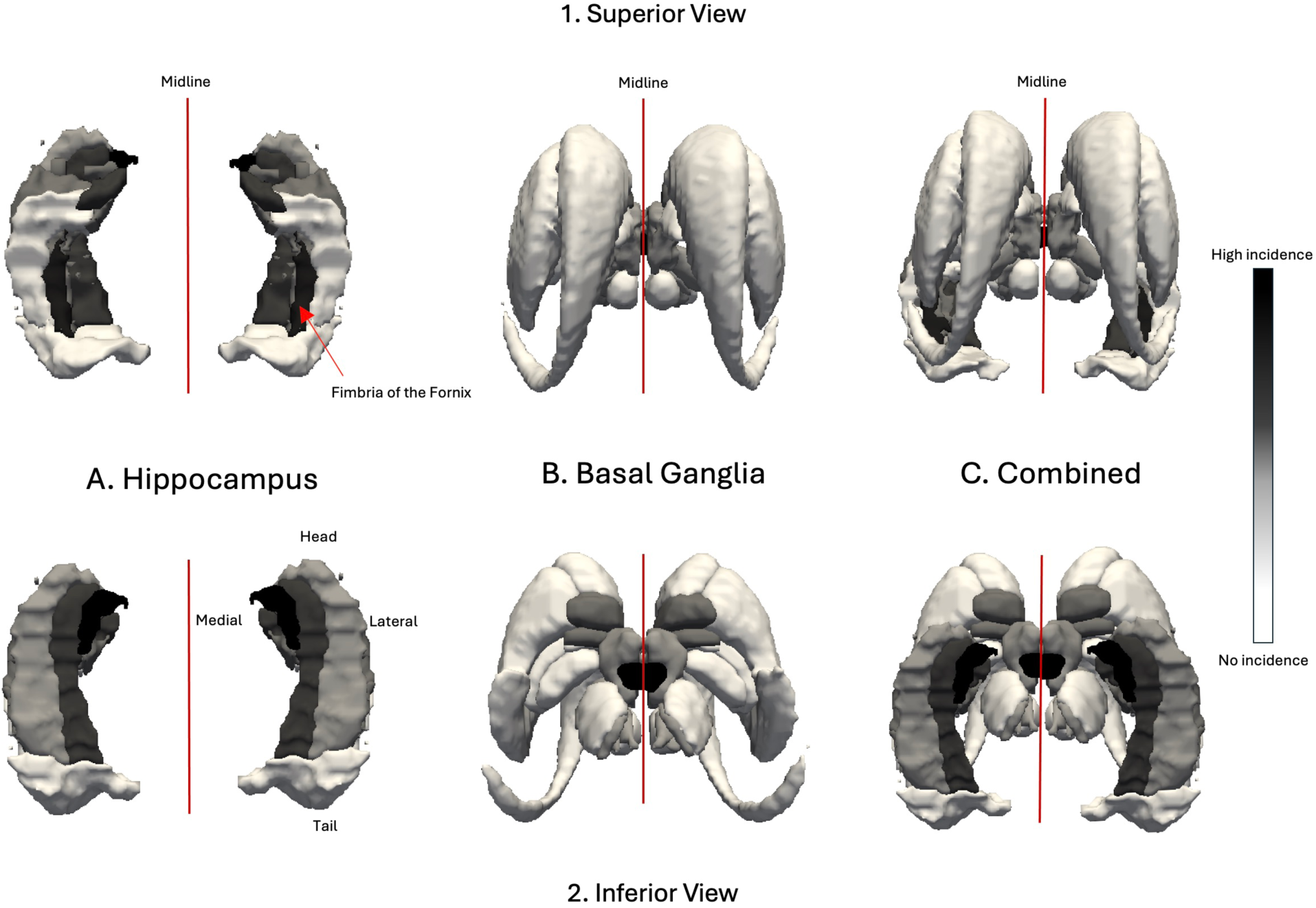
Aggregated regional incidence of abnormal iron markers. This figure presents a 3D model of relative incidence rates aggregated across the “iron-abnormal” mTBI population from a superior (1) and inferior (2) view. Incidence rates are represented in grey scale (high: black; low: white). A. Depicts the hippocampus in isolation. Anteromedial, and commissural (i.e., the fimbria) structures of the more inferior hippocampus show a higher incidence of abnormal z-scores than more superior or lateral structures. The medial aspect of the anterior hippocampal head is more frequently affected than lateral regions or the more distal tail. B. Depicts the basal ganglia in isolation. Substructures of the superiorly-situated basal ganglia show relatively lower rates of abnormal iron markers than the underlying hippocampal formation, with affected regions also primarily located in relatively more inferior and structures near the midline. C. Provides a combined view integrating both hippocampal and basal ganglia regions. The CA1 region has been omitted due to negative z-score.

After subdividing the mTBI participants into iron-normal (20/35; 57%) and iron-abnormal (15/35; 43%) sub-groups based on their individual profiles, a one-way ANOVA showed no significant differences in age between the three groups (iron-normal mTBI, iron-abnormal mTBI, and controls), *F*(2, 32) = 1.12, *p* = 0.3. A Mann-Whitney U test revealed no significant difference in BIST scores between iron-normal (*M* = 35.6, *SD* = 30.8) and iron-abnormal (*M* = 41.7, *SD* = 34.5) mTBI participants, *U* = 120, *p* = 0.5.

### Patterns between symptoms and subcortical iron-related markers

This section is included for descriptive purposes, and relates to observations rather than results of inferential statistics. Elevated iron markers with hippocampal involvement presented alongside self-reported cognitive symptoms (including cognitive impairment, memory deficit, concentration issues, confusion, and clinical notes related to mental fog and bradyphrenia). Disturbances in arousal (fatigue and sleep) and sensory sensitivities (including headache, photophobia, and phonophobia) were reported by 92% of participants with hippocampal abnormalities. Vestibulo-ocular dysfunction (including vestibular symptoms, vertigo, ataxia, and visual disturbance) were slightly less common in this cohort (85%), followed by mood disturbances (77%). Participants with elevated hippocampal iron markers that included the parasubiculum reported cognitive impairment and vestibulo-ocular symptomatology at a 100% incidence rate. Elevated iron markers in the mammillary nucleus, which almost invariably co-occurred with hippocampal regions and rarely with other basal regions, was also associated with high incidences of cognitive symptoms and vestibulo-ocular disturbances, as well as issues related to arousal (86% each). In contrast, relatively lower frequencies of all symptom clusters were reported across participants with abnormal iron markers inclusive of basal ganglia subregions. Cognitive impairment and arousal disturbances occurred in 89% of these participants, vestibulo-ocular and sensory dysfunctions in 78% each, and mood disturbances in 67% (see Table 2 for all symptom/region information).

## Discussion

Group-level analytic methods currently dominate the field of clinical research.^57^ However, the suitability of this approach for comprehensive understanding of neuropathology, particularly in the context of mTBI, may be limited. By focusing on population-level effects, subtle, yet clinically meaningful, inter-individual differences in pathophysiology may be overlooked. To advance the current understanding of deep grey matter iron aggregation in acute mTBI, we conducted the first dedicated individualised analysis of positive susceptibility in subcortical brain regions. Detailed segmentations of the basal ganglia were integrated with comprehensive delineation of the hippocampal subfields; an approach designed to overcome the anatomical limitations of previous group-level QSM investigations. Iron markers across 15 basal ganglia segmentations and 10 hippocampal subfields for individual mTBI participants were assessed relative to a normative control population using z-scores, enabling identification of subcortical regions where iron markers significantly deviated from a healthy reference range.

Results revealed abnormal subcortical iron profiles in approximately half of the mTBI sample, most of which involved at least one hippocampal subfield. Hippocampal abnormalities were not only observationally linked to a high incidence of cognitive symptoms, but were also associated with more frequent occurrence of all symptom clusters relative to participants whose abnormal profiles involved the basal nuclei. Among the hippocampal subregions, the parasubiculum was both the most prevalent ROI to be affected, and universally accompanied self reported cognitive and vestibuloocular symptoms. For iron-abnormal mTBI participants with basal ganglia involvement, elevated iron markers in the mammillary nucleus were also prevalent, occurring at the same incidence rate as markers in the parasubiculum across the entire iron-abnormal cohort. Significant z-scores in the mammillary nucleus most often co-occurred with hippocampal abnormalities or in isolation, rather than concomitant with abnormal z-scores in other basal nuclei. Symptoms related to cognitive, arousal, and vestibulo-ocular dysfunction were the most frequently reported symptoms among mTBI participants with mammillary nucleus involvement. Statistical analysis of injury severity scores between iron-normal and iron-abnormal mTBI participants revealed no significant differences, suggesting that although abnormal subcortical iron profiles may be observationally associated with specific symptom phenotypes, they may not be directly related to injury severity on aggregate.

### Regional injury vulnerability in the hippocampus

The extensive white matter connections between the basal ganglia and both cortical and subcortical structures^87, 88^ increase vulnerability to shear and strain in these regions during mTBI.^10^ This excessive biomechanical loading may also alter iron expression via secondary injury mechanisms.^6^ In addition, the naturally high levels of iron in these nuclei, which under normal conditions support metabolic functions,^6, 14, 65^ may exacerbate risk of trauma-induced cytotoxicity mediated by iron.^89^ However, the higher incidence rates of iron-related abnormalities observed in hippocampal regions compared to the basal ganglia in this individualised study suggest that the prevailing focus on basal structures in the extant literature may represent a critical oversight. Instead, results presented here indicate that the underlying hippocampus, and possibly the surrounding cerebrum,^60^ may instead afford the basal ganglia a relative degree of structural protection from injury (see Fig. 4).

**Fig 4:**
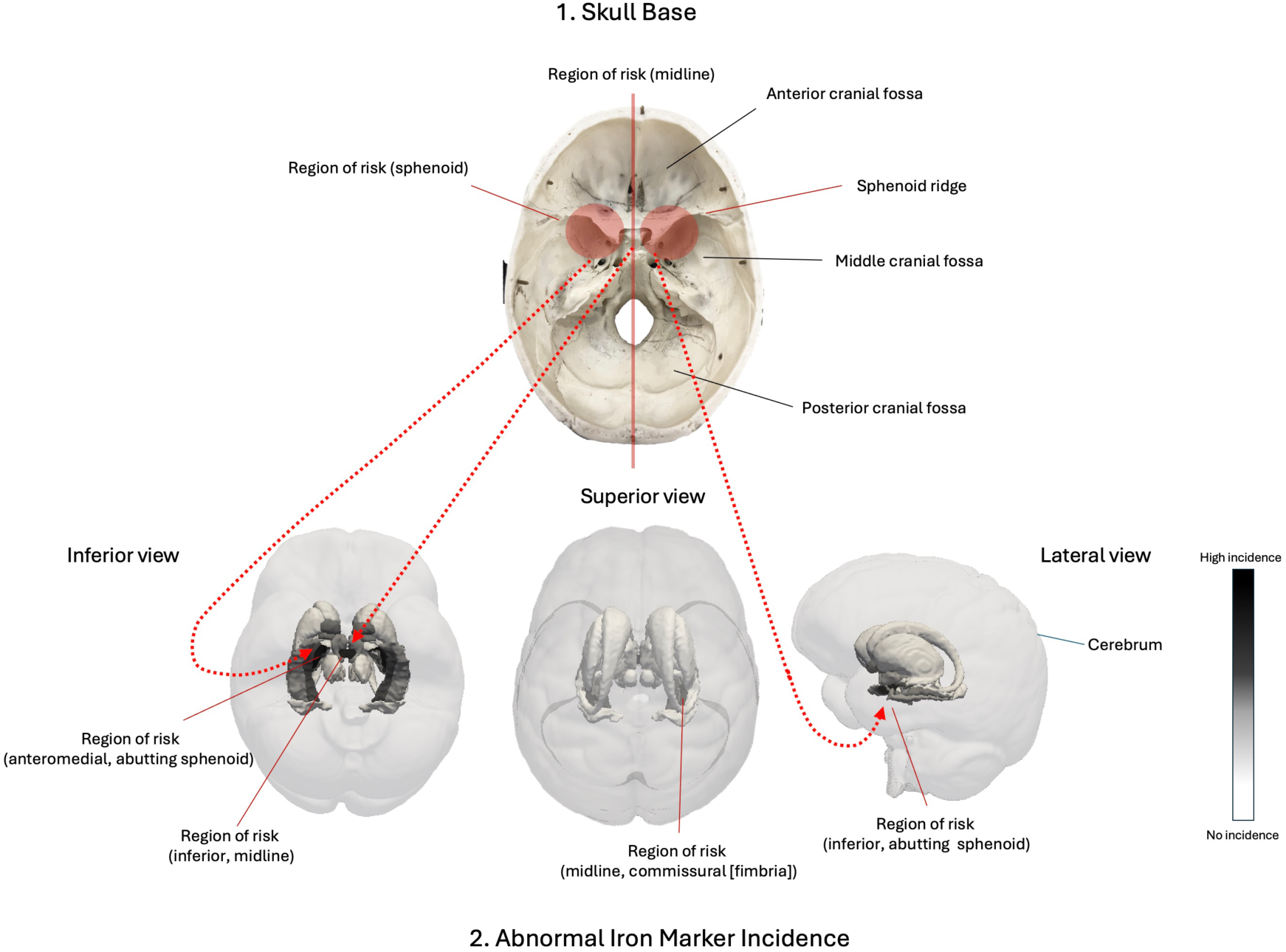
Spatial alignment between regions of heightened injury risk and abnormal iron markers. This figure highlights the anatomical alignment between structural vulnerability and impact biomechanics with aggregate iron markers in subcortical brain regions across “iron-abnormal” mTBI participants. 1. Visualises the cranial base from a superior view. Key regions of mechanical risk, including the sphenoid ridge and midline zones, are highlighted in red. 2. Provides a 3D render showing spatial alignment between regions of risk and incidence rates of abnormal iron markers across the sample. This model provides a visualisation of the vulnerability of inferior structures, particularly in the anterior aspect proximal to the sphenoid ridge, as well as midline and commissural regions, to cranial impact, strain, and force transmission. The basal ganglia, shielded from below by the hippocampus and from above by the cerebrum, may be afforded more protection from injury. The CA1 region has been omitted due to a negative z-score.

Despite the medial temporal lobe’s known vulnerability to traumatic insult,^90, 91^ the hippocampus remains under-studied in QSM-based investigations of mTBI. Viewed in the context of the broader literature, the general predilection for hippocampal iron-related abnormalities across participants in this study indicate local changes to magnetic susceptibility that may be related to iron-driven secondary injury mechanisms^6, 15^ congruent with regional vulnerability of the hippocampus to mechanical deformation in TBI.^91^ Both acute and chronic pathophysiological sequelae associated with hippocampal regions following mild-to-severe traumatic injury can include cytotoxic secondary injury, disruptions to vascular and metabolic function, diffuse axonal injury of afferent and efferent fibers, and even chronic traumatic encephalopathy (CTE)-associated tauopathy and focal atrophy as a downstream pathological event,^90, 92^ for which iron overload may be a mediating factor.^6–8, 93^ On aggregate, the results of the present study suggest that the hippocampus is a primary site of mTBI-related pathophysiological processes.

### Towards a region-of-risk model of mTBI

The distinct, subfield-specific patterns that emerged when abnormal iron markers were aggregated across participants highlights not only the hippocampus but also specific subfields and spatial locations as regions with heightened vulnerability to injury. Injury biomechanics and morphology of the skull base may contribute to increased mechanical loading and strain, focal tissue damage, and a resultant dynamic cascade of secondary injury mechanisms which may include altered iron signalling in select vulnerable, and interconnected, regions (see Fig. 4). Specifically, the concentration of iron markers in the parasubiculum, mammillary nucleus, and fimbria support and extend findings from a previous group-wise cortical QSM investigation^61^ which demonstrated localisation of abnormal positive susceptibility values to the adjacent parahippocampal gyrus.

Anatomically, the head of the hippocampus is situated approximately 2 cm from the irregular sphenoid ridge at the junction of the anterior and middle cranial fossa, and is particularly susceptible to mechanical deformation over this bony protrusion during impact.^94^ The parasubiculum, as part of this region,^95^ may thus be more vulnerable to injury than hippocampal substructures located peripherally to this region of heightened injury risk (e.g., the subiculum) or in the more distal hippocampal tail. This observation is supported by a high degree of spatial overlap between areas of varying injury vulnerability, as informed by cranial-dural architecture and injury biomechanics, and the relative incidence rates of abnormal z-scores observed here which cluster at the anterior aspect of the hippocampus (the hippocampal head), and diminish with proximity to this region (see Fig. 4). These results are also congruent with prior research noting the anterior aspect of the medial temporal lobe as a region of risk.^94, 96^ Data from investigations of temporal lobe epilepsy suggests that parasubicular neurons are hyper-excitable,^97^ which may contribute to iron accumulation as a consequence of excitotoxicity in mTBI,^7^ further exacerbating regional risk. Additionally, the contribution of iron deficiency to neuron loss^14^ provides a plausible explanation for the abnormally low z-score observed in the hippocampal CA1 region of a single participant in this study, reinforcing a hippocampus-specific model of tissue disruption.

Beyond the medial temporal lobe, midline and commissural structures, including the fornix, are increasingly recognised as regions prone to excessive biomechanical loading during sr-mTBI impacts, and have been associated with memory and cognitive impairments in 30-50% of cases and removal from play in 30%.^98^ Damage to midline structures has also been cited as a risk factor for subsequent mTBI,^99^ and although these observations were not directly related to the structures under investigation in the present study, the incidence of abnormal iron markers in the mammillary nucleus and fimbria observed in this sample is nonetheless noteworthy. The fimbria is a key component of the fornix; a commissural tract not only connecting the bilateral hippocampus, but the hippocampus to the mammillary bodies.^94, 100^ Evidence of dyshomeostatic mechanisms focal to this region may be related to head kinematics during impact, whereby rotational forces are a likely cause of strain-related diffuse injury,^101, 102^ which may be particularly damaging to the fornix^98^ and related nuclei. In addition, the mammillary bodies, located on the inferior aspect of the diencephalon, are paired structures that sit on either side of the midline and are connected by the intermammillary sulcus.^103^ These nuclei are also related to the commissural fornix, forming the terminals of its anterior pillars.^104^

Degeneration of both the fornix and mammillary bodies has been extensively documented in TBI, which is attributed to medial temporal lobe trauma and disruption of these regions as downstream targets.^94^ The mammillary bodies receive projections from the parasubiculum,^104^ and abnormal iron markers in this region may represent a more subtle, but pathologically significant, manifestation of degenerative excitotoxic processes that exist on a continuum with more severe TBI. The interconnection of these two regions may not only explain equivalent incidences of iron-related abnormalities in these regions, but their frequent co-occurrence within individual participant results (see Table 2). Taken together, these data not only reinforce the importance of subfield-specific investigations of the hippocampus and granular segmentations of the basal anatomy, but also provide evidence that sites of cellular distress signalling can be mapped to potential injury mechanisms via quantification of positive susceptibility and the use of individualised analytic techniques. This lends support to the utilisation of individualised study designs to detect heterogeneous changes in brain tissue content, and contributes to the current understanding of the relationships between potential iron overload, injury dynamics, and mTBI pathophysiology.

### Regional iron dysregulation, symptom cluster, and Papez circuit integration

Disruption to cellular integrity can impede function, resulting in changes to cognition and behaviour that may be related to specific injury biodynamics and loci of neuronal injury.^93^ Although elevated subcortical iron markers were not associated with greater self-reported symptom *severity*, and causal inference is precluded in this sample, the universal incidence of symptom clusters within the cognitive domain among participants with abnormal hippocampal iron markers is consistent with extant literature linking memory and cognitive deficits to medial temporal lobe damage^90^ and iron dyshomeostasis.^7, 9^ In addition to the general concentration of abnormal iron markers within the hippocampal subfields broadly, the mammillary nucleus and parasubiculum emerged as consistent sites of abnormal iron loading which could speculatively be related to symptom phenotype.

Although the specific role of the parasubiculum in mTBI-induced pathology and associated symptomatology remains under explored, its involvement in key pathways mediating cognitive and vestibulo-ocular functions may offer a plausible explanation for clinical deficits reported by participants presenting with abnormal iron profiles in this region. Participants with elevated iron markers in the mammillary nucleus also reported similar symptoms, with an additional high incidence of arousal disturbances (see Table 2). Functional and structural connectivity between these two regions^104, 105^ may account not only for the overlapping symptomatology, but also for the frequent co-occurrence of abnormal iron markers.

As part of the subicular complex, the parasubiculum occupies a central position in the medial temporal lobe memory system, supporting memory formation.^106, 107^ This is exemplified by observations of pyramidal cell loss and the formation of severe neurofibrillary tangles (NFTs) in the parasubiculum in Alzheimer’s disease (AD)^108^ with memory loss as a central feature.^109^ The parasubiculum also plays a critical role in spatial information processing. ^110, 111^ This region is anatomically and functionally connected to the area prostriata, which projects to visual, motor, and auditory regions,^106^ and interfaces with the entorhinal cortex, a region critical for both memory formation and integration of vestibular and directional cues in spatial navigation.^107, 112^ Directional and spatial orientation signals are further modulated by connections to the mammillary bodies. “Head-direction” cells responsive to head orientation and spatial encoding in the presubicular and parasubicular cortices^110, 111, 113^ transmit signals to the lateral mammillary nucleus to facilitate navigation and orientation.^114^

The mammillary bodies also serve as a central pathway for memory processes. In addition to receiving hippocampal outputs via the fornix,^100^ the mammillary bodies relay these signals to the anterior thalamic nuclei through the mammillothalamic tract.^115^ This relay function is a critical component of the Papez circuit central to emotion and memory.^116^ Damage to this fundamental network has been associated with a variety of neurological disorders that have cognitive impairment as a defining feature, including AD, Parkinson’s disease (PD), semantic dementia, and global amnesia.^116, 117^ Atrophy of the mammillary bodies is also distinguishing feature of Korsakoff syndrome, a degenerative neurological disorder related to chronic alcohol abuse which is characterised by episodic memory deficits,^114^ and linked to dysfunction within Papez circuit.^117^ Within the context of mTBI, pathology of the mammillary bodies has been cited as an antecedent of memory impairment stemming from Papez circuit disconnection in retired athletes with a history of repetitive mTBI.^118^ By extension, it is plausible that similar symptoms reported by participants with evidence of iron accumulation in these key, highly interconnected, regions might arise from dysfunction of these central circuits.

Finally, the high incidence of issues related to arousal also reported by participants with abnormal iron markers in the mammillary nucleus could speculatively be attributed to disruption of histaminergic neurons within the mammillary bodies that regulate arousal and wakefulness. Damage to these neurons has been identified as a mediating factor in sleep dysregulation following mTBI,^119^ corroborating this hypothesis.

### The potential of an individualised approach for enhancing research and clinical care

Collectively, the distribution of abnormal iron markers across subcortical nuclei suggests a region-of-risk model informed by injury biomechanics and a complex interplay with cranial and dural architecture (see Fig. 4). It is well-established that different biomechanical forces give rise to heterogeneous and dynamic injury cascades, complicating the study of mTBI pathophysiology.^93, 120^ The results presented here suggest that characterising dyshomeostatic iron signalling at the individual level may serve as a viable proxy for secondary injury mechanisms occurring in a subset of individuals, and at specific spatial locations. These data indicate that mTBI induces iron overload in at least one subcortical region for 43% of participants with mTBI. Taken together with the lack of identifiable differences in positive susceptibility between mTBI participants and controls in a previous extensive group level investigation^44^ and in the literature more broadly,^30–36^ this finding suggests that iron dyshomeostasis in the deep grey matter may not constitute a feature of mTBI generalisable to all cases. However, the use of individualised modelling likely enables identification of instances at the single-subject level where iron accumulation may be occurring, providing biologically informative results that are otherwise lost to averages. This suggests that targeted interventions, for example heavy metal chelation therapies,^121–125^ may be beneficial for some individuals, emphasising importance of identifying individual variation that could inform the development of future clinical trials and specific treatment strategies.^57^ In addition, leveraging inherent tissue susceptibility variations has the potential to identify not only specific injury patterns but also individuals at elevated risk of suboptimal recovery, subsequent injury, or localised tissue pathology, provided that study designs are suitably responsive. The observed relationships between loci of abnormal iron markers and symptom clusters also alludes to the potential of individualised approaches to elucidate how distinct patterns of cellular disruption may be related to specific clinical phenotypes.

As an extension of the same logic, the relative under representation of abnormal iron markers in the CA4 in this sample, coupled with the observation of decreased negative susceptibility for mTBI participants in this region in prior group-level analyses,^44^ suggests that while the CA4 may be vulnerable to disruption of negative susceptibility sources, mechanisms of iron dyshomeostasis in this region are less prevalent. This, in turn, highlights the utility of group-wise analyses to identify potential signatures of mTBI that are congruent with established disturbance of hilar cell populations in head injury^45–48^ and temporal lobe epilepsy.^49^ These convergent data underscore the importance of tailoring the analytic strategy choices to the research objective.

This work also underscores inherent limitations of the standard methodological focus prevalent in contemporary QSM-based mTBI research. Specifically, identifying hippocampal subfields and the mammillary nucleus as primary loci of acute, iron-related mTBI pathophysiology suggests that prior research concentrated only on major basal ganglia structures,^30–33, 35^ or using gross hippocampal segmentations,^34, 36^ is likely insensitive to discrete regions differentially affected by injury. At minimum, this work highlights the need to incorporate both individualised analyses and detailed segmentations into future research designs to improve understanding of regional, inter-individual tissue content changes following mTBI, and provides evidence that calls for the scientific community embrace new paradigms that account for injury heterogeneity^120^ and more detailed segmentations^6^ are well-founded.

Lastly, the congruence between dominant spatial distributions of abnormal iron markers in this sample and diagnostic features of CTE are concerning given the putative relationship between the two.^68, 126, 127^ In particular, the presence of NFTs in the hippocampus and mammillary bodies is diagnostic of CTE and CTE burden (high versus low).^92^ Identifying these specific regions as common locations of potential iron dyshomeostasis in mTBI raises further questions about the interrelatedness of acute cascades that may present as later-life pathology in a subset of individuals.

### Limitations and future research

There is a lack of any widely established best-practice method to normalise the HC z-distribution and other individualised QSM research has employed less stringent filtering at three times IQR to detect more severe TBI pathology.^53^ Future research would benefit from standardised practices for outlier detection. The mammillary nucleus, in particular, was more extensively filtered than other regions. This may have contributed to a greater proportion of abnormal z-scores in this region, despite a residual sample size that is still more robust than other comparable studies.^53^ In addition, the use of nonparametric approaches for between-group comparisons of iron-normal and iron-abnormal mTBI participants is limited by the inherent challenges associated with small and unequal sample sizes, which may compromise the statistical power and reliability of these findings; thus, the results of the Mann-Whitney U test to investigated differences in injury severity scores should be interpreted with caution.

Natural age-related increases in subcortical iron can also be particularly pronounced within this specific age range,^65^ and although the ages of the HC and mTBI participants were closely matched, some confounding effects may still be introduced. For example, the relative under representation of abnormal iron markers in the basal substructures (with the exception of the mammillary nucleus) may be related to the potential confounding effects of age-related increases in iron. These effects may be particularly relevant in regions known to exhibit elevated iron content during normal ageing such as the red nucleus, substantia nigra, globus pallidus, putamen, and caudate,^62, 63, 65–67^ many of which showed no evidence of abnormal individualised iron markers compared to the HC population. Despite the lack of statistically significant age differences between groups in all relevant analyses, it is still possible that age-related variability in regional iron content may yield an inherently broader normative distribution, thereby confounding identification of abnormal iron markers in the mTBI cohort.

Although this study used detailed segmentations, some structural limitations remain. For instance, the mammillary bodies comprise functionally and structurally distinct subregions. Evidence from murine models suggests that head-direction cells are present in the lateral, but not medial, mammillary nuclei.^114^ Future investigations should consider additional segmentation granularity, whilst balancing the need to detect subregion-specific pathophysiology and challenges related to multiple comparisons. In addition, individualised approaches should integrate additional MRI imaging modalities, for example, to investigate white matter pathway integrity as a marker of axonal injury (e.g., by diffusion MRI) and/or functional connectivity via functional MRI. A multi-modal approach would facilitate a deeper understanding of how specific tracts, networks, and nodes are implicated in injury-related pathophysiology and symptomatology, and how this may relate to patterns of cellular disruption in grey matter on QSM. The use of single-echo data for QSM reconstruction inhibits the application of true intra-voxel magnetic source separation.^128–133^ While intervoxel thresholding has been applied to investigations of AD^81^ and healthy ageing^82^ with meaningful results, this approach does not distinguish the heterogeneous susceptibility sources within voxels, constraining inference at the biological level. Lastly, the identification of relationships between abnormal regional iron markers and symptomatology made here were observational only; the small sample size and some ambiguity in self-reported symptoms precludes the use of inferential statistics. Additional intra-individual overlap in abnormal z-scores in basal and hippocampal ROIs also inhibits precise delineation of the relationship between basal versus hippocampal z-score clusters and clinical symptom phenotype. The reader should bear in mind that any inferences are speculative, and do not represent a one-to-one symptom-to-ROI mapping.

## Conclusions

This study presented the first dedicated individualised QSM investigation of iron-related tissue content changes in subcortical grey matter regions following mTBI. The results highlighted the inter-individual heterogeneity in regional iron markers, and underscored the vulnerability of specific hippocampal subfields congruent with established regions-of-risk in the TBI literature. Injury severity was not significantly different between mTBI participants presenting with abnormal iron markers and those who did not, however, iron-related markers localised to certain hippocampal subfields and the mammillary nucleus appeared to be related to distinct clinical phenotypes. These results support the use of analytic techniques sensitive to inter-individual variation to identify instances of injury-induced grey matter micropathology, which have been largely absent from the group-wise QSM investigations of mTBI that have been conducted to date.

## Author Contributions

**Christi A. Essex** (Conceptualisation, Methodology, Project Administration, Validation, Software, Formal Analysis, Investigation, Resources, Data Curation, Writing - Original Draft, Writing - Review & Editing, Visualisation); **Mayan J. Bedggood** (Writing - Review & Editing, Project administration, Investigation); **Jenna L. Merenstein** (Writing - Review & Editing); **Catherine Morgan** (Methodology, Writing - Review & Editing); **Helen Murray** (Writing - Review & Editing); **Samantha J. Holdsworth** (Writing - Review & Editing); **Richard L.M. Faull** (Writing - Review & Editing); **Patria Hume** (Writing - Review & Editing); **Alice Theadom** (Conceptualisation, Methodology, Writing - Review & Editing, Funding acquisition, Supervision); **Mangor Pedersen**(Conceptualisation, Methodology, Writing - Review & Editing, Funding acquisition, Supervision)

## Data Availability

All data produced in the present study are available upon reasonable request to the authors

## Acknowledgements

We extend thanks to Amabelle Voice-Powell and Cassandra McGregor for their contribution to the data collection, and Tania Ka’ai for bringing her perspective to cultural considerations on this study. In addition, we thank Axis Sports Concussion Clinics, particularly Dr Stephen Kara, for their assistance with recruiting sr-mTBI participants and personnel at the Centre for Advanced Magnetic Resonance Imaging (CAMRI) for their assistance collecting MRI data. We also acknowledge Dr Tim Elliot for radiological reporting of all participants and Siemens Healthineers for the use of a work-in-progress (WIP) prototype sequence for the acquisition data used to perform QSM.

## Data availability

De-identified MRI data and code used for image processing and statistical analysis can be made available upon request to the corresponding author.

